# Motivations underlying co-use of benzodiazepines and opioids in the UK: a qualitative study

**DOI:** 10.1101/2024.11.29.24318190

**Authors:** G Vojt, HE Family, H Poulter, C Bailey, D Cavallo, AP Abdala Sheikh, S Karimi, N Booth, P Da Silva, L Aitken, S Stewart, M Hickman, G Henderson, J Scott, J Kesten

**Author notes:** Corresponding author: Gabriele Vojt. Joint last authors: Jennifer Scott and Joanna Kesten.

## Abstract

**Background:** Drug-related deaths have substantially increased over the past decade in the UK, particularly in Scotland. Co-using opioids and benzodiazepines (prescribed and/or illicit) is a risk factor contributing to rising mortality. This study identified motivations in people’s co-use with the aim of informing prescribing and harm reduction interventions to address drug-related deaths.

**Methods:** We interviewed 48 people who co-use opioids and benzodiazepines and/or z-drugs (zopiclone and zolpidem) in Glasgow (n=28), Teesside (n=10) and Bristol (n=10). Most participants self-identified as male (n=37, 77%), white (n=45, 94%) and had a mean age of 43 years (range: 25-61 years). The majority reported at least one overdose experience, and poor mental health including trauma. Interviews were semi-structured, conducted by an academic and/or peer researcher, and analysed using reflexive thematic analysis.

**Results:** Participants’ motivations for co-using drugs mapped onto two interlinked meta-themes: (1) Functional motivations included co-using to augment drug effects, self-medicate or help to generate income. (2) Experiential motivations described participants’ desires to achieve (a) ‘buzz’ (feeling energised), (b) ‘glow’ (feeling comforted) (c) ‘oblivion’ (escaping trauma and adversity), and (d) feeling ‘gouchy’ (physical and mental sensations of ebbing in and out of glow and oblivion). Those typically seeking a glow or buzz wished to minimise the risks of co-use. Participants aiming to achieve oblivion or feeling ‘gouchy’ felt ambivalent about the risk of overdose.

**Conclusions:** The importance of assessing motivations to co-use should be recognised and routinised as part of harm reduction, (co-)prescribing and medication assisted treatments to reduce mortality risk.

## Introduction

The co-use of benzodiazepines and/or z-drugs (zopiclone and zolpidem) with opioids is associated with increased risk of drug-related-deaths (DRDs) globally (World Health Organisation, 2023). In Scotland, most DRD involve opioids (82%) and benzodiazepines (70%) (Scottish Government [SG], 2023), a characteristic of the ‘Scottish overdose epidemic’ (McAuley et al., 2022). Co-use is defined as the consumption of two substances, either simultaneously or sequentially, resulting in interacting effects determined by duration of drug action (Votaw et al., 2019). Co-use of benzodiazepines and opioids (referred to as co-use hereafter), especially when consumed within a wider polysubstance use pattern (e.g., combined with stimulants such as cocaine, and alcohol) are acute risk factors in opioid toxicity, i.e., fatal and non-fatal overdoses. Therefore, interventions for co- and polysubstance use are urgently required to prevent DRDs and improve the support available to people who co-use. One approach to addressing co-use is the co-prescribing of opioid antagonist treatment (OAT) and benzodiazepines, however, this alone is unlikely to be a sufficient response due to its association with increased mortality (Matheson et al., 2024) and because co- and poly-drug use do not occur in a vacuum. For example, co-use and increased risk of DRDs are associated with lower socio-economic status, poverty (SG, 2022), low rates of healthcare and treatment access, engagement (McAuley et al., 2022) and retention (SDF, 2021), poor physical (Lewer et al., 2022) and mental health (SG, 2022; Mental Welfare Commission, 2022). There is therefore a need to incorporate mental health support into treatment. The feasibility of this is currently being assessed in a trial in Scotland by combining a low-dose benzodiazepine maintenance intervention with OAT, peer support and brief trauma-informed sessions (Berry et al., 2023).

Understanding a person’s needs to (co-)use drugs is key in optimising treatment and service delivery. In Scotland, Medication Assisted Treatment (MAT) standards (SG, 2021) and MAT guidance for benzodiazepine harm reduction emphasise the importance of staff’s ‘understanding [of] presenting issues, predisposing, precipitating and perpetuating factors’ underpinning drug use (Public Health Scotland, 2023, p. 3-4). This is reinforced by the UK clinical management guidance (2017) on working with people who use drugs. Practically this means that all staff supporting people at high risk of DRD are required to psychologically formulate, i.e., hypothesise how and why a person’s substance use occurs, under what circumstances and what factors keep it going (Division of Clinical Psychology, 2011). Here, understanding individuals’ motivations to co-use contributes to considering the person within context, enabling staff to co-develop treatment plans and goals, agreeing on priorities in both the short- and long-term, as well as retaining the person in services by creating personal meaning, hope and agency (Association of Clinical Psychologists, 2022).

Theoretically, motivations to use alcohol and other substances are conceptualised as interactions of people’s expectations, their learned history and traits (e.g., responses to substance use, past reinforcements) and current contextual factors (e.g., affect, availability, coping skills and physical setting) (Cox & Klinger, 2011). The literature on motivations specifically underpinning co-use suggests a broad dichotomous framework consisting of (1) self-therapeutic (removing negative emotional states such as anxiety) and (2) hedonic (or seeking euphoria) motivations (Vogel et al., 2013; Fatseas et al., 2009). However, these findings are anchored in quantitative research designs. A qualitative and mixed methods review of motivations for polydrug use, across different populations, reported eight motivational patterns (Boileau-Falardeau et al., 2022). Drugs were used sequentially to (1) alleviate withdrawal, and (2) prolong euphoria (‘being high’). Drugs were used simultaneously to (3) balance the effects of drugs, (4) counteract the effects of drugs, (5) enhance euphoria, (6) reduce overall drug use and associated harms (especially from alcohol use), and (7) mimic the effects of substances. When studies did not specify the temporal sequence of polysubstance use, this was typically related to motivations to (8) self-medicate (in relation to pain).

In summary, the existing qualitative literature tends to report motivations in functional terms (i.e., self-therapeutic) with less written about hedonic motivations, which may restrict and impact on staff’s understanding of factors influencing co- and poly-drug use. Findings predominantly reflect a US and Canada context, with evidence accumulated between 2010-2015 and outwith a focused ‘benzodiazepine-opioid’ lens. This is a major challenge as misinterpreting motivations, mis- implementing prescription guidance among healthcare professionals and under-prescribing can lead to people illicitly seeking substances to address or prevent withdrawal (e.g., McAuley et al., 2022) and potentially disengaging the person from services.

In this paper, we examine self-reported motivations to co-use benzodiazepines/z-drugs and opioids to inform harm reduction and intervention designs to prevent DRDs. We included z-drugs as these were (1) developed as an alternative to benzodiazepines and (2) there is typically little overlap between people who use benzodiazepines and those who use z-drugs illicitly (Hockenhull et al., 2021).

## Methods

The current paper is part of a research project funded by the Medical Research Council (ref: MR/W029162/1) involving both qualitative and neuropharmacology studies to examine the interactions between benzodiazepines and opioids which may increase the likelihood of fatal overdose.

### Study design

We conducted a qualitative study with people who co-use benzodiazepines/z-drugs and opioids in three diverse cities in the UK - Glasgow (Scotland), Bristol (South-West England); and Teesside (North-East England). We chose these locations to produce a heterogenous understanding of people’s motivations, across diverse socioecological backgrounds and drug cultures. Glasgow has high rates of DRDs with benzodiazepines and opioids, Teesside has high opioids-related DRDs and high rates of z-drug use (Poulter et al., 2023) but less poly-drug use involving benzodiazepines and opioids compared to Glasgow. Bristol has a high prevalence of opioid use, high crack cocaine-opioid poly-drug use and lower than average opioid DRDs.

### Participants

We interviewed 48 people with current and recent (in the past 6 months) co-use of benzodiazepines/z-drugs and opioids, with 28 interviews in Glasgow, ten in Teesside and ten in Bristol. Table 1 summarises demographics for the total sample and by study location.

**Table 1.** Participant demographics for the total sample and per study location.

|  |  | **Glasgow**<br>**(n=28)** | **Bristol**<br>**(n=10)** | **Teesside**<br>**(n=10)** | **Total (n=48)** |
| --- | --- | --- | --- | --- | --- |
| **Gender** | male | 21 (75.0%) | 7 (70.0%) | 9 (90.0%) | 37 (77.1%) |
|  | female | 7 (25.0%) | 3 (30.0%) | 1 (10.0%) | 11 (21.9%) |
| **Mean age (SD)** |  | 42.2 (9.7) | 46.4 (6.2) | 40.7 (7.3) | 42.8 (8.7) |
| **Age range** |  | 25-61 | 41-61 | 30-50 | 25-61 |
| **Ethnicity** | White British | 3 (10.7%) | 5 (50.0%) | 9 (90.0%) | 17 (35.4%) |
|  | White English | -- | 1 (10.0%) | -- | 1 (2.1%) |
|  | White Scottish | 25 (89.3%) | -- | 1 (10.0%) | 26 (54.2%) |
|  | Black British | -- | 1 (10.0%) | -- | 1 (2.1%) |
|  | White African | -- | 1 (10.0%) | -- | 1 (2.1%) |
|  | Other (Norse) | -- | 1 (10.0%) | -- | 1 (2.1%) |
|  | Declined to answer | -- | 1 (10.0%) | -- | 1 (2.1%) |
| **Housing** | Home/houseless | 10 (35.7%) | 4 (40.0%) | 3 (30.0%) | 17 (35.4%) |
|  | Supported accommodation | 2 (7.1%) | -- | 3 (30.0%) | 5 (10.4%) |
|  | Rented/ own accommodation | 16 (57.2%) | 6 (60.0%) | 4 (40.0%) | 26 (54.2%) |
| **Alcohol (self-defined)** | Heavy | 8 (28.6%) | 2 (20.0%) | 2 (20.0%) | 12 (25.0%) |
|  | Moderate | 5 (17.9%) | 3 (30.0%) | 2 (20.0%) | 10 (20.8%) |
|  | Minimal | 7 (25.0%) | 2 (20.0%) | 2 (20.0%) | 11 (22.9%) |
|  | None | 6 (21.4%) | 3 (30.0%) | 4 (40.0%) | 13 (27.1%) |
|  | Undisclosed | 2 (7.1%) | -- | -- | 2 (4.2%) |
| **Non-fatal overdose experience** | Self-reported | 21 (75.0%) | 7 (70.0%) | 10 (100.0%) | 38 (79.2%) |
| **Poor mental health** | Diagnosed and self-reported | 28 (100.0%) | 9 (90.0%) | 8 (80.0%) | 45 (93.8%) |

### Procedure

#### Recruitment

Potential participants were identified and recruited via gatekeepers using the following inclusion criteria: aged 18+ years, able to converse in English and had co-used benzodiazepines or z-drugs and opioids in the past six months. We recruited from drug treatment and harm reduction services, homelessness outreach, residential crisis and stabilisation services, and support groups including a women’s only group. In line with the collaborative and intensive pragmatic qualitative (CLIP-Q) approach (Horwood et al., 2022), peer researchers and staff from these services acted as gatekeepers and supported the research team with access, identification and recruitment of a diverse sample of research participants with a range of co-use experiences. All gatekeepers received a brief bullet-point summary of the study aims, incentives and inclusion criteria. Recruitment posters were also made available for potential participants to directly contact the research team.

Gatekeepers provided verbal information on the study and handed information sheets to potential participants. Those who expressed an interest to take part in an interview contacted the academic researcher or liaised via the gatekeepers. Researchers arranged interviews with potential participants based on their preferences, or when interviews had been pre-arranged by the gatekeeper, the researcher would attend at the time and location preferred by the participant. Opportunistic recruitment also took place with researchers regularly visiting services. Interviews typically took place in secure and staffed settings; on two occasions interviews were conducted in communal settings such as a cafe as per participant preference. All interviewees received £10, in cash or voucher depending on the relevant organisation’s policy as a token of appreciation for participants’ time and help.

#### Data collection

All data collection was conducted in individual interviews by an academic researcher (GV, HF, HP) only, or co-conducted with a trained local peer researcher (n=18 interviews), depending on the peer researchers’ availability and agreement of the participant and host organisation. Interviews took place between November 2022 and September 2023 in-person or by telephone. Peer researchers who were staff or volunteers at recruiting services had lived experience of substance use, were familiar with the local drug culture and were primarily involved to ensure that participants felt comfortable and safe (Roche et al., 2010; Rhodes et al., 2010). They supported data collection and interpretation by probing for details during interviews and providing additional insights informed by their lived experience. All peer researchers attended project- and topic guide specific qualitative training (adapted from Hickman et al., 2019). Interviews lasted a mean of 50mins (SD=19mins), ranging from 20 to 103 min. The median was 48 mins. We anticipated a sample size of at least 45 interviews would be sufficient. The decision to cease collecting data was informed by the principle of information power (Malterud et al., 2016) assessing the data collected in relation to the study aim, sample specificity (participant characteristics relating to the phenomenon under study), quality and depth of the data and planned analyses.

All interviewees provided written informed consent. Throughout all interviews, peer and/or academic researchers sought continuous consent and ensured interviewees were aware of their right not to answer questions. Debrief and, where required, emotional support was provided after each interview. Peer researchers and/or the peer research organisation received payment for their time. Interviews were audio recorded, transcribed (intelligent verbatim) and anonymised.

#### Reflexivity

Using the CLIP-Q approach, we wrote summaries and reflexive notes after each interview to inform future interviews. We continuously reflected and considered findings within weekly team meetings to facilitate translation and triangulation between study sites and teams. The credibility, integrity and applicability of findings were checked with peer researchers (with lived experience of co-use) and expert stakeholders in each study location at the analysis stage.

#### Patient and Public Involvement and Engagement (PPIE)

Collaborators and service users were involved in all stages of the research development and design. We initially held a workshop in Bristol with a diverse group of people who co-use to gather input on the design, content and language used in the recruitment materials and interview topic guide, including review for appropriate local terminology. We also utilised PPIE and stakeholder input and reality-checking in the interpretation of our findings, via two online workshops.

#### Conceptual frameworks

The socioecological framework (McLeroy et al., 1988) was the conceptual basis for the study and the design of the topic guide. Through the socioecological framework lens, we considered the motivations and wider characteristics of co-using drugs within the context of sociological, cultural, economic structures. We also used this framework when considering the implications of our findings at the intra- and interpersonal, organisational and system level (combining community, policies and systems) (see figure 1).

**Figure 1.**
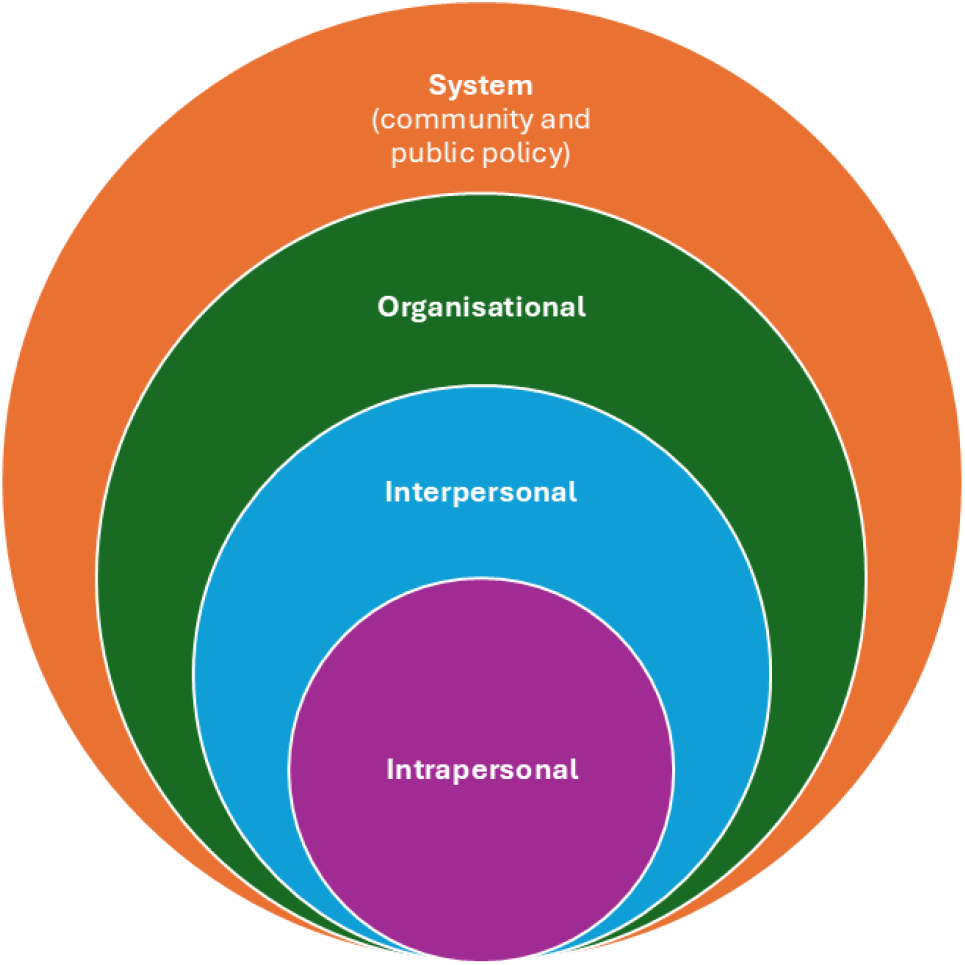
The socioecological framework adapted to health promotion impacts.

### Topic Guide

The topic guide explored why, when and how people co-use within individual, interpersonal, organisational and system/policy structures. The topic guide focussed on the interviewee’s experience of co-using benzodiazepines/z-drugs along with opioids; motivations and patterns of co-use; the role of different types of benzodiazepines and opioids in non-fatal overdose experiences; how the risks of co-use were managed and the characteristics of valuable interventions. Interviews also captured socio- demographic characteristics, drug treatment history, mental and physical health conditions. Images of illicit benzodiazepines, z-drugs and opioids submitted to WEDINOS (see https://www.wedinos.org/) for drug testing and locally generated testing data were also used as prompts. Patterns of co-use (Family et al., 2024) and perceptions of co-use risks and harm reduction (Poulter et al., 2025) are reported separately.

### Data analysis

All transcripts were analysed inductively and deductively using reflexive thematic analysis (Braun & Clarke, 2019) in NVivo and on paper. We took a data driven approach following Braun and Clarke’s five-phased approach: (1) all anonymised interview transcripts were read and re-read to aid data familiarisation, (2) we open-coded transcripts, guided by participants’ meanings associated with motivations. Next, (3) we collated these codes into themes, (4) we reviewed the generated themes deductively, using existing literature to finally (5) define and name them. We noted that final themes clustered into two higher informational groups, and therefore defined these as meta-themes. Data analysis was led by a team of researchers (GV, HF, HP); all themes and findings were discussed and refined with senior researchers (GH, JS, JK), and with the wider research team (CB, DC, AS, MH ).

## Ethics approval

Ethical approval for this study was obtained from the Faculty of Health Sciences Committee for Research Ethics, University of Bristol (ref 11906).

## Results

We identified motivations to co-use benzodiazepines or z-drugs with opioids, mapping onto two broad meta-themes; ‘functional motivations’ and ‘experiential motivations’, with associated themes and sub- themes. Reflecting the underlying principles of motivational theory, participants held multiple motivations at the same time, these were dynamic and changed depending on situation, context, mental health state and the person’s expectations from co-use.

We present our findings comparatively by functional and experiential motivations. Figure 2 outlines the themes and subthemes across functional and experiential motivations when co-using benzodiazepines/z-drugs and opioids.

**Figure 2.**
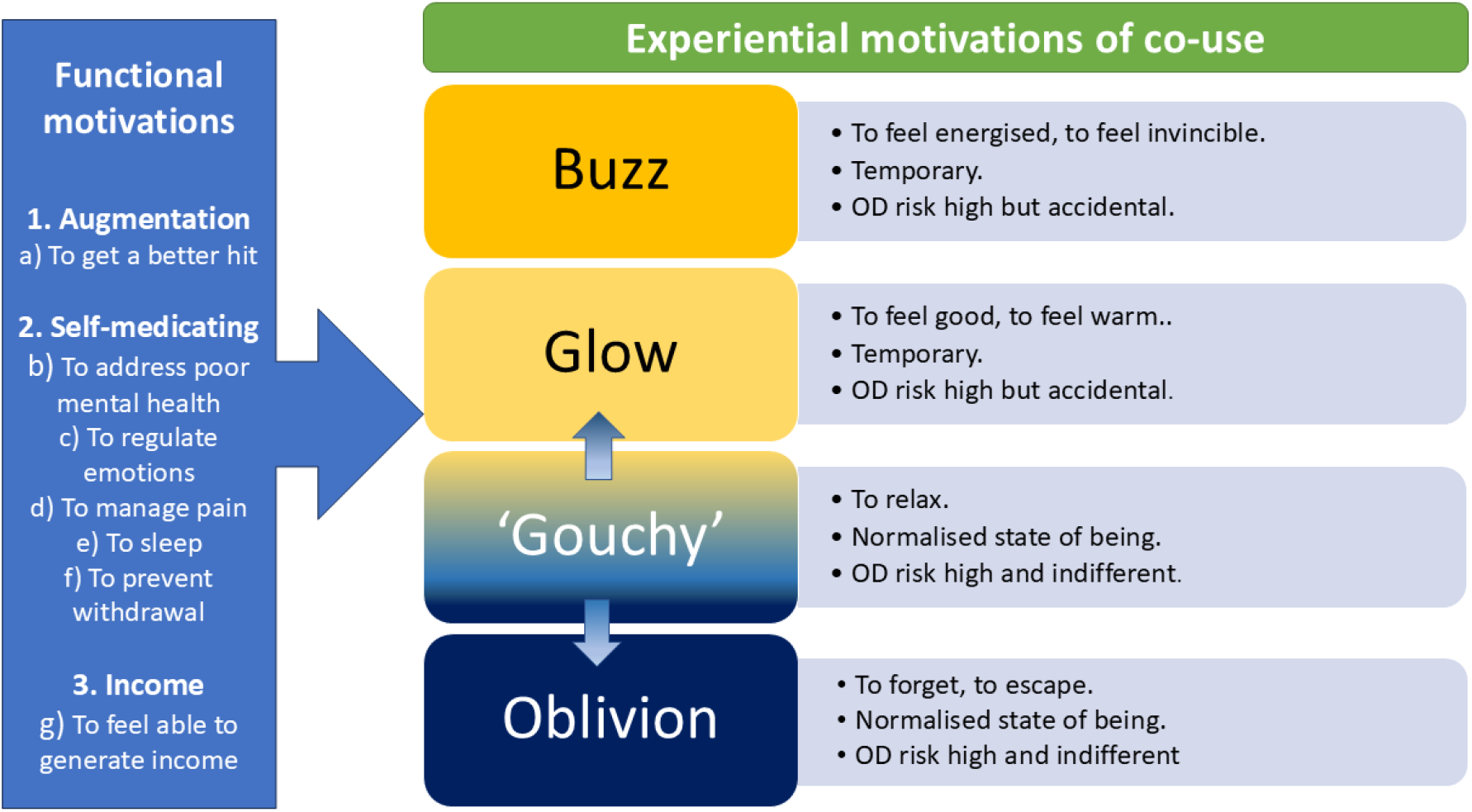
Functional and experiential motivations of co-using benzodiazepines/z-drugs and opioids.

Nb. Many interviewees were uncertain about the actual active ingredients in the substances they used, which prevented linking specific drugs to specific experiences with certainty (McAuley et al., 2022).

Among our sample, co-use of benzodiazepines/z-drugs and opioids ranged from exclusively focussing on benzodiazepines or z-drugs with opioids to co-using within the context of wider poly-drug and alcohol use. Here, participants emphasised one or a set of preferred drugs, often benzodiazepines, z-drugs, heroin or crack cocaine. Participants’ narratives included intentional and accidental, non-planned co-use, and reflected a range of self-monitored, restricted vs intense and binge-like co-use patterns (Family et al., 2024).

## Functional motivations

There were three themes in participants’ functional motivations; these were 1. *‘augmentation’* – to enhance the psychoactive effects of drugs, 2. *‘self-medicating’* – to alleviate poor mental and/or physical health, and 3. *‘income’* – to feel able to generate income. With input from our peer researchers, we established that functional motivations are the primary drivers for co- and poly- substance use.

### 1. Augmentation

Most participants described augmenting, i.e., enhancing the effect (‘hit’ or ‘dunt’) with benzodiazepines or z-drugs with opioids, typically heroin, methadone, oxycodone and codeine. Participants explained that the tolerance to benzodiazepines/z-drugs increases rapidly, therefore co- or poly-drug use was often described as essential to augment benzodiazepine-effects to meet expectations and needs.

> *‘If I was going out to get the both of them* [heroin and street Valium] *I would take some of the street Valium. I would probably take 25 before I went home.* […] *I would take them and then I would go home and smoke some heroin a bit at a time and bring them* [street Valium] *on.*’ (P13, male, Glasgow)

Benzodiazepines/ z-drugs were also co-used to counterbalance the effects of other drugs, e.g., to ‘come down’ from cocaine. While those on maintenance prescription of Buvidal^®^ (buprenorphine slow-release injection) tended to poly-use with benzodiazepines/ z-drugs, they did not consider this as co-use with opioids per se, but rather as separate drug using behaviours that operated in parallel.

### 1. Self-medicating

Here, participants contextualised their co-use as part of being able to function on a daily basis. Sub- themes centre around motivations *‘to alleviate poor mental health’* (e.g., diagnosed anxiety, depression, post-traumatic stress disorder and trauma (particularly among women)), *‘to regulate emotions’* (e.g., stress, social anxiety, daily worries), *‘to induce and enable sleep’*, *‘to manage pain’*, and *‘to prevent withdrawal’*.

Participants often co-used within the context of poor mental health support, insufficient or absent benzodiazepines prescribing and experiences of stigma when asking for support and increases in prescriptions. As a consequence, illicit benzodiazepines in particular were used *‘to alleviate poor mental health’* and *‘to regulate emotions’*. The following participant focuses on the effects of benzodiazepines when self-medicating with the aim of feeling ‘normal’ as opposed to mentally unwell (e.g., paranoid).

> ‘[Benzos] *make me feel good* […] *and make me feel calm, make me feel lucid, make me feel normal in a sense, you know what I mean.’* [Can you say a wee bit more, what does it feel like to be normal?] *‘It just feels calm, less paranoid, feeling like you’re able to, I don’t know, just actually step out the door in a sense, know what I mean. Usually, I just feel like sitting in my room all day, depressed* […] *but when I take my benzos I can go out and I can do things. It’s like I’ve had to come for blood tests today, I’ve did that, know what I mean.’* (P34, male, Glasgow)

> [What is it like to use benzos and heroin together?] *‘Well, I wouldn’t say it’s good, if you know what I mean, because it’s not. It feels like it just blocks your pain out and you’ve just got that numb feeling* […] *It just takes things out your way. And it makes you want to clean the house, want to do stuff, want to go out. Instead of feeling all that scared way, you know.’* (P45, female, Glasgow)

Poor mental health and trauma experiences were also related to the subtheme *‘to induce and enable sleep’* as participants outlined co-use to subdue racing thoughts, primarily achieved by using benzodiazepines and/or z-drugs.

> *‘I think I use drugs not to feel good off them but to try and keep it under control, my mental health. Basically, to block it out because it’s just a daily burden, every day.* […] *Your mind races on a night and you can’t stop it. And your body’s tensed up, so the Zopiclone is to knock me out and the* [street] *Diazepam is to relax me.’* (P25, male, Teesside).Some participants described co-using specifically ‘*to manage pain’*. This participant describes being on prescribed morphine and valium to manage physical pain and discomfort. When his morphine and valium prescription was reduced and in the absence of required professional support, he self-medicated by obtaining street opioids and benzodiazepines (e.g., OxyNorm and pregabalin) on top of his prescriptions.

> *‘I was on 800 of zomorph* [morphine] *a day, so I’ve come down to 110, but I started on 800mls a day and two bottles of Oramorph* [morphine] *a week. Then I came down to* [area]*, they reduced me straight down to 25mgs of valium from 100. I had about 30 mini-strokes, no joke.* […] *I phone my doctor, I said, ‘Listen, I’m in so much pain. I need something for breakthrough pain, like a bottle of oramorph, you’ve reduced me too fast. He said,* ‘[Name], *I’m not your–‘ I wish I could have recorded it, he goes, ‘I’m not your fucking joint dealer, go and buy some smack,’ and hung up.*’ (P17, male, Bristol)

In the sub-theme *‘to prevent withdrawal symptoms’*, the fear or anticipation of potentially withdrawing motivated co-use as much as the actual experience of withdrawal. This then created a cycle of co-use necessitating a regular supply of substances.

> *‘There would be really bad withdrawal symptoms that you would get from them* [co-using street valium and heroin] *so I think all I wanted to do was to get that off as well. It was like a mind thing as well. If you knew you didn’t have them and you knew you had run out it would be a really horrible feeling so you would need to get your heroin and you would need to get your street Valium just to take that edge off it. It was that really horrible feeling you would get from both of them.* […] [If I was withdrawn] *I would basically be feeling absolutely awful. I would be feeling terrible. I would be shaky. I would be just all over the place.’* (P13, male, Glasgow)

While participants tended to describe withdrawal symptoms from benzodiazepines/z-drugs as more severe and debilitating than opioid withdrawal, we cannot determine with certainty in our data whether participants took benzodiazepines/z-drugs specifically to suppress opioid rather than benzodiazepine/z-drug withdrawal.

### 2. Income

Some participants described co-using *‘to feel able to generate income’*, such as criminal activities, sex work and other forms of paid labour. Often, participants’ narratives highlighted a sense of routinised and normalised co-using procedures as part of working and making money.

Co-using to partake in criminal activities (e.g., shoplifting, burglary, selling drugs) served the function of numbing yet energising the person by instilling confidence. Participants also reported that benzodiazepines and z-drugs made them feel invisible and invincible.

> *‘Downers* [heroin] *just block things out. Just what you feel* [is] *numb… and I think see, that pairs well with Valium, like I’m saying I might think I’m alright but other people are seeing me present and I’m mad with it, but I’m not thinking I’m mad with it and when I come back and that whole criminal side, that shit again, when you’re taking Valium you think nobody sees you, you think you’re invisible, you’ve got that confidence for you, you don’t care about anything, and I’m back in shops shoplifting and all that.’* (P39, female, Glasgow)

> ***‘****I used to always get the jail because I used to do robberies. I used to rob post offices and building societies and that, and they* [street Valium] *gave me Dutch courage. The Zopiclone and the Valium, they’d be for Dutch courage. Aye, they make you invincible and good for graft.’* (P1, male, Teesside)

Participants rarely volunteered information specifically on co-using within the context of sex work. Instead, participants – typically females – discussed co-using as a means to do *‘whatever I had to do’* in order to generate enough income to secure further drugs, which then helped them to function in other life areas. Some participants described long-term co-use as a driver to perform professional duties. Often, this was interlinked to people discussing co-use as a means to stay calm and to negate physical symptoms of withdrawal, pain or stress. The following extract exemplifies using illicit benzodiazepines and heroin before engaging in labour jobs, with the focus on heroin as the main drug to function professionally.

> ‘*There were times when I would get up in the morning and I knew I had the job to go to at ten and I would nip away and get myself something, just maybe a little bag of heroin.* […] *I wouldn’t take as many* [street] *benzos but would take a little bit of heroin and that sort of stuff just so I wasn’t shaky. I would do laminate flooring for people and build bits of furniture for them. It wasn’t any major jobs or anything like that but just your handyman jobs that I would do. That got me through that to be honest with you.’* (P13, male, Glasgow)

Similarly, another participant described long-term co-use of heroin (and later methadone), cocaine and illicit benzodiazepines in addition to large quantities of alcohol on a daily basis while working in a managerial position.

> *‘Booze has always been part of my life. It’s always been there. It’s always been a crutch* […] *fuck, I could easily… I could easily drink 24 cans a day, easily, from ten o’clock in the morning, without thinking about it twice* […] *I used to work and hide it* [alcohol use]*. The same as with my drug-taking* [heroin, cocaine and benzodiazepines] *I could work and hide it, and I could still make my work and naebody would know any difference.’* (P29, male, Glasgow)

In summary, across the functional motivations, co-using benzodiazepines/z-drugs and opioids served as a survival mode within a wider context of adverse life events, loss, disadvantage and trauma. For some participants, benzodiazepines/z-drugs were the main substance they needed to activate themselves in the morning, to feel able to function, and therefore any activity thereafter was benefitting from the benzodiazepines intake.

## Experiential motivations

Interlinked to functional motivations but often discussed separately, participants described their motivations to co-use in terms of achieving specific bodily and mental experiences. Experiential motivations described the following themes: 4. *‘buzz’* (feeling invincible and energised), 5. *‘glow’* (feeling comforted), 6. feeling *‘gouchy’* (physical and mental sensations of ebbing in and out of glow and oblivion) and 7. *‘oblivion’* (forgetting or escaping adversity and trauma). Participants in our sample described experiences of *‘buzz’* and *‘glow’* using interchangeable terminology, e.g., both experiences were referred to as relaxing but also energising, depending on dosage, co- or poly- substance use combinations and situational context.

### 3. ‘Buzz’

Those participants who primarily sought to experience a *‘buzz’* described this as a temporary feeling of energy, increased confidence and self-efficacy (a person’s belief in their ability to complete a task or achieve a goal) to the extent that participants believed themselves to be invincible and/or invisible. Seeking this experience was often linked to functional motivations such as engaging in criminal activities, although equally to obtain sufficient energy to perform daily life functions such as doing housework.

> *‘Zopiclone, they work relatively fast. But the effect they have on me* […] *they give me a bit of a high. If I can’t be bothered to do nothing I take a couple of them, get my housework done, and then you just sleep. I’m just ready to shower and then sleep, but sometimes I don’t get that far.’* (P28, female, Bristol)

### 4. ‘Glow’

When defining ‘glow’, participants tended to highlight that these positive experiences were benzodiazepine/z-drug-driven (depending on study location, i.e., zopiclone was primarily discussed in Teesside and not at all in Glasgow) and induced happiness and feelings of safety. Some participants contextualised *‘glow’* by referring to media presentations (e.g., ‘Ready Brek’ porridge advert depicting an animated person with an orange glow while feeling relaxed and content) or describing personal experiences of safety (e.g., when visiting their grandparents and being cared for).

> [What does glow feel like?]*‘The feeling is a very overwhelming feeling of warmth, a feeling of there’s not a person who wouldn’t accept you, it makes me feel good.’* (P48, male, Glasgow)

### 5. ‘Gouchy’

Feeling *‘gouchy’* was thought of as a slow pattern of ebbing in and out of deep relaxation with intermittent sensations of ‘buzz’ or ‘glow’. Opioids were used as the foundation to induce deep relaxation to the extent of being immobile and oblivious to one’s surroundings, with sufficient benzodiazepines consumption to extend this state of relaxation. These two participants describe the changes in experiencing benzodiazepines when re-dosing while gouching.

> *‘When you gouch, your body’s jelly. Your body’s just like jelly. Sometimes you cannae even move.* […] [After using heroin] *you just take about 25 of the valium, shove them in my mouth and chew them, and as I chew, I’ll eat a bit of cake, drink coffee, then we’ll take another 25 and then another 25 as long as we feel great, we feel fine. But we don’t sleep. We gouch, wake up; gouch, we wake up.’* (P21, female, Glasgow)

> *‘It’s* [gouching] *like a peak, it’s like it peaks, it goes down then peaks again and then goes down and then peaks again and then goes down.’* (P43, male, Glasgow)

### 6. ‘Oblivion’

Similar to feeling ‘gouchy’, we identified a motivational theme *‘oblivion’*. The following quote contextualises oblivion within the context of a binge-using pattern; while the participant outlines consuming alcohol, prescribed methadone and diazepam, this occurred within a wider context of poly drug use (e.g., street diazepam, cocaine, pregabalin) depending on financial resources.

> *‘They* [benzodiazepines] *took me into oblivion, they took me away from all my problems, I didn’t need to worry about anything, nothing bothered me, it was just like excuse the line but it was just basically, “Fuck everybody, I can just do what I want and that’s it.” That’s why I liked it because I could just take them and it was like there was no worries, there was no hassle, there was no wondering who was going to phone me today or who’s going to be moaning at me, work isn’t going to be chasing me up all of that. It just went out the window plus into the bargain it sort of gave me confidence, it gave me false confidence, false sort of bravado, all that sort of stuff* […] *I was that much in oblivion with drugs and alcohol combined on top of 100 and odd methadone prescription a methadone prescription a diazepam prescription.’* (P10, male, Glasgow)

Those who co-used to experience oblivion sought a space where they may exist but not necessarily experience or feel. Often, this was associated with avoiding or escaping trauma experiences and memories.

## Overdose risk perception within motivations to co-use

The majority of participants had either experienced a drug overdose or periods of unconsciousness following co-use. Within the motivational themes *‘glow’* and *‘buzz’*, there was a sense that participants perceived their overdose risk as high. Participants acknowledged the likely dangers of using illicit benzodiazepines, however, in the absence of professional support and prescribed benzodiazepines, participants were willing to balance this risk against functional motivations, e.g., to prevent withdrawal. In the experiential motivation themes *‘oblivion’* and *‘gouchy’*, there was also an understanding of the increased risk of overdosing, especially when participants reported co-use in binges or within a wider poly-drug use pattern. However, participants expressed ambivalence with regards to drug related harms. The following two quotes illustrate (1) the lack of concern for the self, and (2) the contrast in priorities and needs between the ‘sober’ self and the ‘emotional’ self.

> *‘I overdosed about eight times in the space of five months but I got to the point where I didn’t care anymore. That’s probably what I was trying to look for the words I was trying to find, the right words when I said I drank to oblivion, I used drugs to oblivion, I just didn’t care* […] *whether I died or I didn’t. As long as I was out of my trolley and out of my face I really didn’t care. I didn’t think, “What if you die when you take this?” I just thought, “I’m taking this to get wasted.” Then come midday I’d be waking up in the* [hospital]*, “What’s wrong?” “You overdosed and you were taking fits, taking seizures.” I’d be like, “Right okay, I’m just going to go now.”’* (P10, male, Glasgow)

> [When you co-use and add alcohol, do you ever worry about your health?] *‘When I’m full of them* [benzodiazepines]*, I don’t know. Like seriously, sober me right now, I’m petrified of death, I am. I don’t want to die. I want to live. I don’t want to exist because that’s like, I feel like my life right now I’m taking the correct steps to do things, but I feel like I’m stagnant and I just exist on the planet and not actually living a life. And I want to live. But when my emotions* [trauma based] *get too much for me to deal with, I turn, don’t care what’s going to happen to me as long as I’m mad with it and I don’t remember.’* (P38, female, Glasgow)

## Discussion

In this paper, we identified inter-related functional and experiential motivations for co-using benzodiazepines/z-drugs and opioids. Functional motivations included a person’s objective to function on a daily basis. Co-use served to ‘self-medicate’, e.g., alleviate/prevent withdrawal and poor mental health, but also to ‘enable income generation’ and to enhance the effects of other drugs (‘augmentation’). Experiential motivations described how co-use felt to the person, cognitively and physically, and ranged from feeling energised (‘buzz’), happy (‘glow’) to deeply relaxed and numb (‘gouchy’ and ‘oblivion’). Overdose risk perceptions tended to be tied in with motivational themes, e.g., ‘buzz’ and ‘glow’ were characterised by being aware of, and wishing to reduce, overdose risk.

Those seeking ‘oblivion’ and feeling ‘gouchy’ tended to express ambivalence about overdose risk and outcomes. While these motivational themes were generated from the total sample; we found no geographical differences when reflecting on location.

### Comparison with existing research knowledge

Our findings extend the existing literature on motivations to co- and poly-substance use at a dichotomous level (self-therapeutic to ‘functional’ vs. hedonistic to ‘experiential’) (Vogel et al., 2013) and at a granular level (Votaw & Witkiewitz, 2021; Boileau-Falardeau et al., 2022). The experiences ‘glow’, ‘oblivion’ and ‘buzz’ sought when co-using build on existing descriptions such as ‘a little glow all round me’ (May et al., 2020), seeking a ‘blackout’ (Fatseas et al., 2009) and wanting to feel ‘euphoric’ (Park et al., 2021). Co-use enabling people to generate income via criminal activities and sex work is well documented (e.g., Votaw & Witkiewitz, 2021). However, our findings further evidence that co-use may also enable people to function physically and mentally in managerial and labour jobs. Our findings on overdose risk perception and awareness by experiential motivations also reflect differences reported across the literature (Votaw & Witkiewitz, 2021; Fatseas et al.,2009; Motta-Ochoa et al., 2017; Neira-Leon et al., 2006; Rowe et al., 2016; Stein et al., 2017; Strang et al., 1999). Central to many of the functional motivations for co-use is self-treatment in line with drugs’ medicinal function (Votaw & Witkiewitz, 2021; Boileau-Falardeau et al., 2022). For example, participants spoke of seeking medication for physical and mental health conditions suggesting unmet treatment needs. Reported challenges in accessing prescribed medications are associated with individuals attempting to address such unmet needs with illicit medications (May et al., 2020). That is, some participants in our study sought out street benzodiazepines to numb the impact of historical and ongoing disadvantage including sexual, violent and psychological trauma, self-reported poor mental health including diagnoses, and challenging life experiences from early childhood to adulthood (e.g., homelessness, poor physical health).

### Implications

Our findings have multiple implications for therapeutic interventions highlighting the need for pathways to screen, diagnose and adequately treat mental health symptoms (e.g., anxiety) and to address the experiential and functional effects of benzodiazepines used by people co-using opioids (McHugh et al., 2017, Vogel et al., 2013). Importantly, understanding motivations for co- and poly- substance use should be routinely assessed and incorporated into clinical decision making on (co-) prescribing, drug treatment interventions and harm reduction. Using the sociological framework, implications can be mapped at the intrapersonal, interpersonal, organisational and system level.

At the **intrapersonal level**, our findings can optimise staff understanding of the importance of identifying motivations in conversation (e.g., during motivational interviewing) with patients. Harm reduction staff in needle and syringe programmes, safer consumption rooms, overdose prevention programmes and policies (Hawk et al., 2017) are likely the key professionals in contact with people who co-use drugs and who may not have access to drug treatment. To strengthen staff understandings, routinised assessments of motivations should be part of clinical and psychosocial assessments. The key is to understand why a person uses drugs, how they started using drugs and what perpetuates the co- and/or poly-use of drugs. By contextualising the person and their motivations to co-use drugs, the therapeutic alliance can be strengthened, and interventions can be kept flexible and relevant to the person co-using drugs. The systematic measurement of motivations for co-use using tools such as an inventory of drug taking situations (Turner, Annis & Sklar, 1997) or a visualised pathway map could help staff to support people with tailored advice, treatment and underpin shared decision making to (co-) prescribe. Such a situation or context specific inventory/ map aligns with motivational theory, i.e., motivations change depending on context, situational and interpersonal expectations. For example, our results demonstrate that benzodiazepine-intake was motivated by both, to get a buzz but also to relax, depending on situation. The experience and expectation of these sensations were related to co-use patterns, dosage and frequency of benzodiazepine/z-drug use (Family et al., 2024).

At the **interpersonal level**, service users’ perceptions and understanding of overdose and drug related harms may require upskilling people who co- and poly-drug use prophylactically to balance individuals’ motivations against risks of harm. This is particularly important considering that motivations to use drugs are not discreet (Votaw & Witkiewitz, 2021; Boileau-Falardeau et al., 2022) nor stable (Vogel et al., 2013). Therefore, prescribers, drug treatment, harm reduction and support staff have the opportunity to help the patient to identify the importance, function and priority co-use has on a daily basis, and more generally across the person’s life. Here, harm reduction staff are encouraged to maximise on all opportunities to engage and assess people’s motivations to co-use to integrate this into a structured referral system. For example, if co-use is primarily driven by functional motivations such as self-medicating to suppress poor mental health, then harm reduction staff can signpost and refer on to specialist services including mental health, counselling and/or drug treatment teams. In terms of experiential motivations, if service users share they primarily co- or poly-use to gouch out or achieve oblivion, then this is likely to indicate a (temporary) crisis situation where the service user is not necessarily focussed on their safety and wellbeing while using drugs. In the short term, the aim then is to ensure the person is supplied with naloxone, and if available be referred to a supervised healthcare setting such as a safer consumption room. In the long term, medication assisted treatment, peer support and psychosocial interventions are essential. Likewise, prescribers’ decision making will benefit from systematically assessing motivational drivers. For example, if a patient describes their motivations to co-use in functional terms, then prescribers could implement split- dosage prescribing tailored to the daily contexts when functional motivations are most prevalent (e.g., early mornings to get out of bed, or later in the day to leave the house to go to the pharmacy). If this is not possible (e.g., due to supervised medication schemes), then prescribers could use motivational understandings to consider changes in the person’s OAT prescription (dosage or type of OAT) and/or manage potential side effects from other prescribed drugs which the person might try to alleviate via co-use. For example, people who co-use benzodiazepines and methadone are reported to have objectively poor sleep and suffer from insomnia (Peles et al., 2009). Prescribers and patients could explore a move from methadone to buprenorphine or consider sleep treatment to circumvent the need to co-use. In this way then, realistic, sustainable and individualised harm reduction and intervention plans can be *co-produced* and implemented in the short- and long-term.

**Organisationally**, a staff culture underpinned by psychologically framed values and ethos should be strengthened. Awareness and an understanding of functional and experiential motivations to co-use should be reinforced. While organisational change in healthcare settings is notoriously challenging (Correa et al., 2020), an example from the UK, Scotland, highlights that healthcare, drug treatment staff and prescribers have been effectively trained up in psychologically formulating people’s motivations to (co-) use drugs (Traynor, personal communication, 2024). Therefore, staff-wide upskilling to consider people’s behaviour within a motivational and psychological context is feasible. However, improved economic resources and funding avenues are required to endorse and implement staff training, in particular to prevent staff burnout when faced with the complex realities and reasons of people’s co-use and life experiences (SDF, 2022). Carlisle and colleagues (2023) underline the dilemma staff in drug treatment services face in terms of organisational policies and pressures. On one hand, staff are required to facilitate successful, drug-free discharges, yet with relatively little time to understand the ‘reasons’ (or motivations) for people’s drug use. Therefore, awareness of our motivational themes and their behavioural impact may support aims to retain service users and help to design response care and treatment planning. This in turn may increase intervention effectiveness in the long term (by being able to update treatment to individual needs and priorities) and reduce potential healthcare and service costs incurred through overdose reactive care and discharge from services (SDF, 2022; Black, 2021). To achieve this, our findings may encourage the establishment of a shared vocabulary between patients and staff, opening up a safe space where potential power differentials can be navigated. In particular, a local/regional list of key terms and associated motivations (e.g., ‘oblivion’) could be used to alert staff to probe further in a non-stigmatising, inclusive and open-ended format (e.g., ‘what is wrong with you?’ vs ‘what has happened to you?’; see Johnstone, 2022). This could then inform referral decisions to appropriate treatments, e.g., trauma, mental health counselling or peer-facilitated activities and support groups.

At the **system** level, the main implications are three-fold: firstly, existing interventions and approaches such as motivational interviewing should be aligned with guidance and psychology formulation principles among staff working with people who co- and poly-drug use. Secondly, structural support provision to reduce the risk of overdose should be endorsed via drug testing, naloxone availability and overdose prevention centres. Thirdly, the underlying motivation for people to co- or poly-substance use is to function, often in disadvantaged, impoverished and unstable contexts. Increasing people’s ability to function, to have a sense of control and agency in their decision-making and daily life means that contributing key social and environmental factors need to be addressed. Therefore, improved housing, employability, social connectivity, interventions to improve wellbeing and engagement with services remain a key aim (Black, 2021) yet a system-wide gap.

## Future research

To optimise treatment and interventions, future gender-responsive research on the trauma and mental health needs are required among people who co-use specifically to self-medicate and/or seek oblivion or to feel gouchy. Further, research exploring service provider views on risks and benefits inherent in current clinical practice and harm reduction approaches and experiences of co-prescribing in the context of opioids and benzodiazepines alongside OAT is needed.

## Strengths and Limitations

We focused on a snapshot of people’s self-reported experiences of co-use, which is unlikely to fully account for the dynamic interactions between motivations, situations and interpersonal responses. We did not include quantitative measures or cross-validation of drug strength or dosage. Further, while our sample was reflective of the national demographics of high-risk groups co-using substances in the UK, we failed to recruit from diverse ethnic backgrounds. We focussed on participants in urban inner cities, and therefore our implications are unlikely to transfer easily to people who co-use in rural settings. We did not examine the motivations and impacts for poly-substance use per se, or for drugs other than benzodiazepines/z-drugs and opioids. Strengths include the involvement of local peer co-researchers and input from a multi-disciplinary team (pharmacists, experimental laboratory, third sector, psychology, behavioural sciences, epidemiology). Analytic generalisations were built through rigorous inductive analysis and triangulation across three geographically diverse research sites to develop broad theories, or conceptualisations in relation to the study aims, objectives and research questions. To support the transferability of the research, we involved peer researchers and national expert stakeholders (including third sector, policy makers, academic researchers and public health professionals) to sense check, fine tune and consider the applicability of implications to other settings and populations.

## Conclusion

This study generated an in-depth and nuanced insight highlighting the complexities and relationships between co-use motivations. Understanding motivations is key to providing individually tailored harm reduction, which in turn link theory to practice and underpin effective treatment and patient centred care. Utilising the understanding of interrelated motivations to co-use should be incorporated into staff training to provide psychosocial support to people in treatment and people benefitting from harm reduction advice and advocacy. Co-prescribing benzodiazepines in the absence of understanding motivations to co-use drugs is likely to meet people’s needs only in the short-term and only for a specific section of people stabilised on OAT and intent on wishing to avoid withdrawals.

## Data Availability

All data produced in the present study are available on application at the University of Bristol data repository. Data access is restricted to bona fide researchers for ethically approved research and subject to approval by the University's Data Access Committee.

## Acknowledgements

We would like to thank the people who took part in this research, the staff and services who helped us to recruit in Glasgow, Teesside and Bristol, and the peer co-researchers Joanna Green, Chris Shilvock, Jade Ritchie, Louise Aitken, Nick Booth and Peter Da Silva who conducted valuable field work for and with us. Thanks also to the expert stakeholders who have acted as ‘critical friends’, reviewed the findings and given their feedback on the relevance for future policy and practice. The views expressed are those of the authors and not necessarily those of the Medical Research Council, the NIHR, the Department of Health and Social Care, or UKHSA.

## References

1. Association of Clinical Psychologists UK (2022). Team formulation key considerations in mental health services. https://acpuk.org.uk/team-formulation-key-considerations-in-mental-health-services/. Accessed August 11, 2024.

2. Berry, K., Matheson, C., Schofield, J., Dumbrell, J., Parkes, T., Hill, D., Kilonzo, M., MacLennan, G., Stewart, D., Ritchie, T., & Turner, M. A. (2023). Development of an intervention to manage benzodiazepine dependence and high-risk use in the context of escalating drug related deaths in Scotland: an application of the MRC framework. BMC Health Services Research, 23(1). 10.1186/s12913-023-10201-7.

3. Black, C. (2021). Review of drugs part two: prevention, treatment and recovery. Independent report. https://www.gov.uk/government/publications/review-of-drugs-phase-two-report/review-of-drugs-part-two-prevention-treatment-and-recovery#foreword. Accessed August 11, 2024.

4. Boileau-Falardeau, M., Contreras, G., Gariépy, G., & Laprise, C. (2022). Patterns and motivations of polysubstance use: a rapid review of the qualitative evidence. Health Promotion and Chronic Disease Prevention in Canada, 42(2), 47–59. 10.24095/hpcdp.42.2.01.

5. Braun, V., & Clarke, V. (2019). Reflecting on Reflexive Thematic Analysis. *Qualitative Research in Sport*, Exercise and Health, 11(4), 589–597. 10.1080/2159676X.2019.1628806.

6. Carlisle, V. R., Maynard, O. M., Bagnall, D., Hickman, M., Shorrock, J., Thomas, K., & Kesten, J. (2023). Should I Stay or Should I Go? A Qualitative Exploration of Stigma and Other Factors Influencing Opioid Agonist Treatment Journeys. International Journal of Environmental Research and Public Health, 20(2), 1526. doi: 10.3390/ijerph20021526.

7. Clinical Guidelines on Drug Misuse and Dependence Update 2017 Independent Expert Working Group (2017). Drug misuse and dependence: UK guidelines on clinical management. London: Department of Health.

8. Correa, V. C., Lugo-Agudelo, L. H., Aguirre-Acevedo, D. C., Contreras, J. A. P., Borrero, A. M. P., Patiño-Lugo, D. F., & Valencia, D. A. C. (2020). Individual, health system, and contextual barriers and facilitators for the implementation of clinical practice guidelines: a systematic metareview. Health Research Policy and Systems, 18(1). https://health-policy-systems.biomedcentral.com/articles/10.1186/s12961-020-00588-8

9. Cox, W. M., & Klinger, E. (2011). A motivational model of alcohol use: Determinants of use and change. In W. M. Cox & E. Klinger (Eds.), Handbook of motivational counseling: Goal-based approaches to assessment and intervention with addiction and other problems (2nd ed., pp. 131–158). Wiley Blackwell.

10. Division of Clinical Psychology (2011). Good practice guidelines on the use of psychological formulation. Leicester: Division of Clinical Psychology, British Psychological Society. http://shop.bps.org.uk/good-practice-guidelines-on-the-use-of-psychologicalformulation.html. Accessed August 11, 2024.

11. Family, H., Vojt, G., Poulter, H., Bailey, C.P., Abdala Sheikh, A.P., Cavallo, D., Karimi, S., Booth, N., Da Silva, P., Aitken, L., Stewart, S., Hickman, M., Henderson, G., Scott, J. & Kesten, J.M. (2024). A qualitative study of Benzodiazepine/Z-drug and Opioid co-use patterns and overdose risk. medRxiv 2024.07.26.24311053. https;//doi.org/10.1101/2024.07.26.24311053.

12. Fatséas, M., Lavie, E., Denis, C., & Auriacombe, M. (2009). Self-perceived motivation for benzodiazepine use and behavior related to benzodiazepine use among opiate-dependent patients. Journal of Substance Abuse Treatment, 37(4), 407–411. 10.1016/j.jsat.2009.03.006

13. Hawk, M., Coulter, R. W. S., Egan, J. E., Fisk, S., Reuel Friedman, M., Tula, M., & Kinsky, S. (2017). Harm Reduction Principles for Healthcare Settings. Harm Reduction Journal, 14(1). 10.1186/s12954-017-0196-4.

14. Hickman, M., Dillon, J.F., Elliott, L., De Angelis, D., Vickerman, P., Foster, G., Donnan, P., Eriksen, A., Flowers, P., Goldberg, D., Hollingworth, W., Ijaz, S., Liddell, D., Mandal, S., Martin, N., Beer, L.J.Z., Drysdale, K., Fraser, H., Glass, R., Graham, L., Gunson, R.N., Hamilton, E., Harris, H., Harris, M., Harris, R., Heinsbroek, E., Hope, V., Horwood, J., Inglis, S.K., Innes, H., Lane, A., Meadows, J., McAuley, A., Metcalfe, C., Migchelsen, S., Murray, A., Myring, G., Palmateer, N.E., Presanis, A., Radley, A., Ramsay, M., Samartsidis, P., Simmons, R., Sinka, K., Vojt, G., Ward, Z., Whiteley, D., Yeung, A. & Hutchinson, S.J. (2019). Evaluating the population impact of hepatitis C direct acting antiviral treatment as prevention for people who inject drugs (EPIToPe) - a natural experiment (protocol). BMJ Open, 9(9), Article e029538. 10.1136/bmjopen-2019-029538.

15. Hockenhull, J., Black, J. C., Haynes, C. M., Rockhill, K., Dargan, P. I., Dart, R. C., & Wood, D. M. (2020). Nonmedical use of benzodiazepines and Z-drugs in the UK. British Journal of Clinical Pharmacology, 87(4), 1676–1683. 10.1111/bcp.14397.

16. Horwood, J., Pithara, C., Lorenc, A., Kesten, J. M., Murphy, M., Turner, A., Farr, M., Banks, J., Redwood, S., Lambert, H., Donovan, J. L., & NIHR ARC West Behavioural and Qualitative Science Team. (2022). The experience of conducting collaborative and intensive pragmatic qualitative (CLIP-Q) research to support rapid public health and healthcare innovation. Frontiers in Sociology, 7, 970333. 10.3389/fsoc.2022.970333.

17. Johnstone, L. (2022). Straight talking introduction to psychiatric diagnosis (second edition). Pccs Books.

18. Lewer, D., Brothers, T. D., Hest, N. V., Hickman, M., Holland, A., Padmanathan, P., & Zaninotto, P. (2022). Causes of death among people who used illicit opioids in England, 2001–18: a matched cohort study. The Lancet Public Health, 7(2), e126–e135. 10.1016/S2468-2667(21)00254-1.

19. Malterud, K., Siersma, V. D., & Guassora, A. D. (2016). Sample Size in Qualitative Interview studies: Guided by Information Power. Qualitative Health Research, 26(13), 1753–1760. 10.1177/1049732315617444.

20. Matheson, C., Vucic, C., Dumbrell, J., Robertson, R., Ritchie, T.; Duncan, C., Kessavalou, K., Woolston, C. & Schofield, J. (2024). Clinical Outcomes of Benzodiazepine Prescribing for People Receiving Opioid Agonist Treatment: A Systematic Review of the Evidence. Pharmacy, 12(5), 152. 10.3390/pharmacy12050152.

21. May, T., Holloway, K., Buhociu, M., & Hills, R. (2020). Not what the doctor ordered: Motivations for nonmedical prescription drug use among people who use illegal drugs. International Journal of Drug Policy, 82, 102823. 10.1016/j.drugpo.2020.102823.

22. McAuley, A., Matheson, C., & Robertson, J. (2022). From the clinic to the street: the changing role of benzodiazepines in the Scottish overdose epidemic. International Journal of Drug Policy, 100, 103512. 10.1016/j.drugpo.2021.103512.

23. McHugh, R. K., Votaw, V. R., Bogunovic, O., Karakula, S. L., Griffin, M. L., & Weiss, R. D. (2017). Anxiety sensitivity and nonmedical benzodiazepine use among adults with opioid use disorder. Addictive Behaviors, 65, 283–288. 10.1016/j.addbeh.2016.08.020.

24. McLeroy, K. R., Bibeau, D., Steckler, A., & Glanz, K. (1988). An Ecological Perspective on Health Promotion Programs. Health Education Quarterly, 15(4), 351–377. 10.1177/109019818801500401.

25. Mental Welfare Commission for Scotland (2022). Ending the Exclusion: Care, treatment and support for people with mental ill health and problem substance use in Scotland. https://www.mwcscot.org.uk/sites/default/files/2022-09/EndingTheExclusion_September2022.pdf. Accessed August 11, 2024.

26. Motta-Ochoa, R., Bertrand, K., Arruda, N., Jutras-Aswad, D., & Roy, É. (2017). “I love having benzos after my coke shot”: The use of psychotropic medication among cocaine users in downtown Montreal. International Journal of Drug Policy, 49, 15–23. 10.1016/j.drugpo.2017.07.012.

27. Neira-León, M., Barrio, G., Brugal, M. T., de la Fuente, L., Ballesta, R., Bravo, M. J., Silva, T. C., & Rodríguez-Martos, A. (2006). Do Young Heroin Users in Madrid, Barcelona and Seville have Sufficient Knowledge of the Risk Factors for Unintentional Opioid Overdose? Journal of Urban Health, 83(3), 477–496. 10.1007/s11524-006-9054-5.

28. Park, T. W., Sikov, J., dellaBitta, V., Saitz, R., Walley, A. Y., & Drainoni, M.-L. (2021). “It could potentially be dangerous… but nothing else has seemed to help me.”: Patient and clinician perspectives on benzodiazepine use in opioid agonist treatment. Journal of Substance Abuse Treatment, 131, 108455. 10.1016/j.jsat.2021.108455.

29. Peles, E., Schreiber, S., & Adelson, M. (2009). Documented poor sleep among methadone-maintained patients is associated with chronic pain and benzodiazepine abuse, but not with methadone dose. European Neuropsychopharmacology, 19(8), 581–588. 10.1016/j.euroneuro.2009.04.001.

30. Poulter, H. L., Walker, T., Ahmed, D., Moore, H. J., Riley, F., Towl, G., & Harris, M. (2023). More than just “free heroin”: Caring whilst navigating constraint in the delivery of diamorphine assisted treatment. International Journal of Drug Policy, 116, 104025. 10.1016/j.drugpo.2023.104025.

31. Public Health Scotland (2023). MAT standards informed response to benzodiazepine harm reduction guidance. https://publichealthscotland.scot/media/24430/mat-standards-informed-response-for-benzodiazepine-harm-reduction-guidance-december2023.pdf. Accessed August 11, 2024.

32. Rhodes, S. D., Malow, R. M., & Jolly, C. (2010). Community-Based Participatory Research: A New and Not-So-New Approach to HIV/AIDS Prevention, Care, and Treatment. AIDS Education and Prevention, 22(3), 173–183. 10.1521/aeap.2010.22.3.173.

33. Roche, B., Guta, A. & Flicker, S. (2010). Peer research in action I: models of practice. Toronto, (ON).

34. Rowe, C., Santos, G.-M., Behar, E., & Coffin, P. O. (2016). Correlates of overdose risk perception among illicit opioid users. Drug and Alcohol Dependence, 159, 234–239. 10.1016/j.drugalcdep.2015.12.018.

35. Scottish Drugs Forum (2021). Drug-related deaths in Scotland: MSP Briefing. https://sdf.org.uk/wp-content/uploads/2024/05/Drug-related-deaths-in-Scotland-MSP-Briefing.pdf. Accessed August 11, 2024.

36. Scottish Drugs Forum (2022). Identifying & preventing burnout in frontline services for people who use drugs & alcohol. www.sdf.org.uk/resources/. Accessed August 11, 2024.

37. Scottish Government (2021). Medication Assisted Treatment (MAT) Standards for Scotland: Access, Choice, Support. https://www.gov.scot/publications/medication-assisted-treatment-mat-standards- scotland-access-choice-support/. Accessed August 11, 2024.

38. Scottish Government (2022). National Mission on Drug Deaths: Plan 2022-2026. https://www.gov.scot/publications/national-drugs-mission-plan-2022-2026/pages/3/. Accessed August 11, 2024.

39. Stein, M. D., Anderson, B. J., Kenney, S. R., & Bailey, G. L. (2017). Beliefs about the consequences of using benzodiazepines among persons with opioid use disorder. Journal of Substance Abuse Treatment, 77, 67–71. 10.1016/j.jsat.2017.03.002.

40. Strang, J., Griffiths, P., Powis, B., Fountain, J., Williamson, S. & Gossop, M. (1999). Which drugs cause overdose among opiate misusers? Study of personal and witnessed overdoses. Drug and Alcohol Review, 18(3), 253–261. 10.1080/09595239996383.

41. Turner, N. E., Annis, H. M., & Sklar, S. M. (1997). Measurement of antecedents to drug and alcohol use: Psychometric properties of the Inventory of Drug-Taking Situations (IDTS). Behaviour Research and Therapy, 35(5), 465–483. 10.1016/s0005-7967(96)00119-2.

42. Vogel, M., Knöpfli, B., Schmid, O., Prica, M., Strasser, J., Prieto, L., Wiesbeck, G. A., & Dürsteler- MacFarland, K. M. (2013). Treatment or “high”: Benzodiazepine use in patients on injectable heroin or oral opioids. Addictive Behaviors, 38(10), 2477–2484. 10.1016/j.addbeh.2013.05.008.

43. Votaw, V. R., Geyer, R., Rieselbach, M. M., & McHugh, R. K. (2019). The epidemiology of benzodiazepine misuse: A systematic review. Drug and Alcohol Dependence, 200, 95–114. 10.1016/j.drugalcdep.2019.02.033.

44. Votaw, V. R., & Witkiewitz, K. (2021). Motives for Substance Use in Daily Life: A Systematic Review of Studies Using Ecological Momentary Assessment. Clinical Psychological Science, 9(4), 216770262097861. 10.1177/2167702620978614.

45. World Health Organization (2023). Opioid overdose. World Health Organization. https://www.who.int/news-room/fact-sheets/detail/opioid-overdose. Accessed August 11, 2024.

